# New-Onset Heart Failure in Patients with Obesity using Tirzepatide or Semaglutide

**DOI:** 10.64898/2026.09.21.26363546

**Authors:** Mohammed Al-Nusair, Tala A. Haddad, Shriya Khurana, Jowan Al-Nusair

**Affiliations:** Department of Internal Medicine, Georgetown University- MedStar Washington Hospital Center, 110 Irving St NW, Washington, DC 20010, USA; H. Lee Moffitt Cancer Center, 12902 USF Magnolia Drive, Tampa, FL 33612, USA

**Keywords:** Tirzepatide, Semaglutide, Heart Failure, GLP-1, Obesity

## Abstract

**Background:** Tirzepatide and semaglutide improve functional status and heart failure (HF) events in patients with obesity and HF with preserved ejection fraction. Tirzepatide has been associated with greater control of cardiometabolic risk factors in patients with obesity compared to semaglutide. Whether tirzepatide is associated with lower risk of new-onset HF than semaglutide in patients with obesity is underexplored.

**Methods:** A retrospective cohort study was conducted using data pooled from the TriNetX platform. Patients with obesity who initiated therapy with tirzepatide or semaglutide during the period Jun 4, 2021 to Aug 13, 2026 were identified and 1:1 propensity-score matched to produce two study cohorts. The one-year incidence of new-onset HF was calculated and compared between the two cohorts. Secondary outcomes included new-onset systolic HF, new-onset diastolic heart failure, new-onset atrial fibrillation or flutter, all-cause mortality, and hospital encounters. Cox proportional hazards models were fitted to calculate hazard ratios (HR) along with 95% confidence intervals (CI).

**Results:** Two well-matched cohorts (N=424,269 in each) were formed. The incidence of new-onset HF was lower in the tirzepatide group (1.0% vs. 1.3%; absolute risk difference −0.3%, 95% CI −0.35 to −0.26; HR 0.9, 95% CI 0.86─0.93). The tirzepatide group were also at lower risk of all-cause mortality (HR, 95% CI: 0.8, 0.77─0.91) and new-onset systolic HF (0.8, 0.79─0.92). All other secondary outcomes occurred at similar rates in both study groups.

**Conclusion:** Among patients with obesity, tirzepatide therapy was associated with lower risk of new-onset HF compared to semaglutide although the effect sizes were modest.

## INTRODUCTION

Obesity has emerged as an important chronic noncommunicable disease, with an estimated prevalence of 40%, that is projected to increase by 20% over the next two decades [15,28]. It is associated with an elevated risk of cardiovascular disease including heart failure (HF), coronary artery disease and stroke [18]. Similarly, HF has become a common cause of morbidity and mortality among American adults, with a prevalence that is expected to rise from 6.7 million to 11.4 million in 2050, and a lifetime risk of one in four people [36].

Major risk factors for HF have traditionally included coronary artery disease, hypertension and diabetes. However, the increasing prevalence of obesity has attracted interest in investigating its role in the pathogenesis of HF [3]. Obesity is commonly found among patients with HF [5]. Around two thirds of participants in the National Health and Nutrition Examination Survey, 2015-2018, with HF also self-reported a diagnosis in obesity [7]. Obesity is also associated with an increased risk of incident HF, with a stronger relationship with HF with preserved ejection fraction (HFpEF).[14,29] For every one-unit increase in body mass index (BMI), the risk of HF is estimated to rise by 5-7% [17]. The population attributable fraction (PAF) for obesity is higher in HFpEF compared to HF with reduced ejection fraction [9]. Obesity is thought to underlie a distinct pathophysiological phenotype of HFpEF [5,26], through direct harmful effects on myocardial structure and function, mediated by systemic inflammation, neurohormonal pathway activation, and autonomic dysregulation. Obesity is also related with poorer quality of life and more symptoms in patients with HF,[5,30] along with more frequent HF hospitalizations [18,34].

Lifestyle interventions including dietary modification and physical exercise have traditionally been the primary method for weight loss and are associated with improved cardiovascular outcomes [12], as well as improved functional capacity and symptom burden in patients with HFpEF [13]. Their efficacy is modest however, at around 5-10% weight loss [11], and difficult to sustain over a longer period of time [18,35]. Earlier anti-obesity medications were limited by concerns over safety and minimal efficacy [22,23]. The advent of incretin mimetics have ushered in a new era of anti-obesity pharmacotherapy. These agents offer greater weight loss than lifestyle modifications while exposing the patient to less risk than bariatric surgery, and they have been demonstrated to improve cardiovascular outcomes and HF hospitalizations in patients with HFpEF [6,18–20,27].

The STEP-HFpEF trials randomized obese patients with or without type 2 diabetes (T2D), who had HF with an ejection fraction of 45%, to semaglutide, a glucagon-like 1-receptor agonist (GLP1-RA), or placebo, and demonstrated improved quality of life functional status measures including the Kansas City Cardiomyopathy Questionnaire, six-minute walk test, and New York Heart Association functional class [6,19,20]. In the SUMMIT trial, patients with obesity and HFpEF were randomized to take tirzepatide, a dual glucose-dependent insulinotropic polypeptide (GIP) and GLP1-RA, or placebo, and demonstrated beyond the STEP-HFpEF program, a reduction in HF events in the tirzepatide arm, although event numbers were modest [27]. A recent retrospective cohort study demonstrated a reduced incidence of new-onset HF (NOHF) in patients with obesity who were using tirzepatide therapy compared to matched controls [37].

Tirzepatide has been demonstrated to achieve greater weight reduction and glycemic control compared to monoagonists of GLP-1 receptors [2,4,32]. In the SURPASS-2 trial [10], tirzepatide was associated with greater diabetic control and weight loss compared to semaglutide in patients with T2D. Similarly, in the SURMOUNT-5 trial of patients with obesity without T2D, therapy with tirzepatide lead to greater weight and glycemic control compared to semaglutide [2]. Whether tirzepatide is associated with lower risk of incident HF compared to semaglutide in patients with obesity is underexplored.

The current study aimed to compare the five-year risk of NOHF between obese patients treated with tirzepatide or semaglutide using a retrospective cohort design.

## METHODS

### Study Setting and Design

The current study employed a retrospective cohort design using real-word de-identified data collected by accessing the TriNetX platform. The TriNetX Collaborative Research Network pools data from the electronic health records (EHR) of 170 healthcare organizations across North America, South America, Europe, the Middle East, Africa, and Asia Pacific. The TriNeX platform collects data from various healthcare settings including inpatient admissions, emergency department (ED) visits, outpatient clinics, and electronic health encounters. Available data include demographics (age, sex, race, ethnicity), diagnoses (using the International Statistical Classification of Diseases and Related Health Problems [ICD] − 10 codes), procedures (Current Procedural Terminology [CPT] codes), laboratory results, medications prescribed, and clinical outcomes.

### Study Population

The present study collected data on adult patients, older than 18-years, with a diagnosis in obesity (ICD-10 E66), who had initiated therapy with tirzepatide or semaglutide on or after June 4, 2021, the date on which semaglutide received approval from the US Food and Drug Administration (FDA) for the management of obesity, and until August 13, 2026. The exposure group was defined by the initiation of tirzepatide after a diagnosis in obesity. The control group was defined by the initiation of semaglutide after obesity diagnosis. The starting date of data collection or index date was defined by the first prescription of tirzepatide or semaglutide after a documented diagnosis of obesity. Patients were followed until the occurrence of an outcome event, death, loss to follow-up, five-years (end of follow-up period), or July 5, 2026 (end of study period). Patients who had prescriptions for a GLP1-RA or dual GIP/GLP1-RA other than the one defining the index event within six months from the index event were excluded. Other exclusion criteria included the presence of medical contraindications to the tirzepatide or semaglutide as detailed in the drug packaging; namely, medullary thyroid cancer (ICD-10: C73) and multiple endocrine neoplasia type 2 (ICD-10: E31.22, E31.23).

### Outcomes

The current study compared the five-year risk of two primary outcomes, NOHF and all-cause mortality between the two study groups. New-onset heart failure was defined as the occurrence of a first diagnosis in HF (ICD-10: I50). Secondary outcomes included new-onset systolic HF (ICD-10: I50.2 or I50.4); new-onset diastolic HF (ICD-10: I50.3); new-onset atrial fibrillation or flutter (ICD-10: I48); and any hospital encounter (ED visit or hospital admission).

### Data collection and definitions

One-to-one propensity score matching of clinically relevant covariates was performed to adjust for possible confounding factors. Covariates included in propensity score matching were sociodemographic factors (age at index, sex, race, ethnicity, socioeconomic status), comorbidities (hypertension, dyslipidemia, ischemic heart disease, atrial fibrillation or flutter, valvular heart disease, cerebrovascular disease, peripheral arterial disease, sleep apnea, chronic kidney disease, and chronic obstructive pulmonary disease), lifestyle factors (tobacco smoking, alcohol use disorder), body mass index, systolic blood pressure, medications (insulin, metformin, sodium-glucose cotransporter-2 [SGLT2] inhibitors, beta-blockers, renin-angiotensin-aldosterone system inhibitors, mineralocorticoid receptor antagonists), and laboratory values (low-density lipoprotein cholesterol, glycated hemoglobin [HbA1C], and estimated glomerular filtration rate). All covariates were assessed beginning one year prior to the index event. Covariate definitions are reported in **Table S1** in the Supplementary Materials.

### Ethics approval and informed consent

All data collected in the current study were extracted from the TriNetX platform, which only carries de-identified patient data per the de-identification standard defined in Section §164.514(a) of the HIPAA Privacy Rule. Accordingly, the current study was exempt from institutional review board (IRB) approval and patient consent. The study was performed in line with the principles of the Declaration of Helsinki. All data analyses are secondary analyses of existing data. The study does not involve intervention or interaction with human subjects.

### Statistical analysis

Sociodemographic and clinical characteristics were documented and compared between the two study groups at baseline. Categorical variables were reported as numbers and percentages and continuous variables as medians and interquartile ranges (IQR). Propensity sore matching was carried out by fitting logistic regression models, that were adjusted for all covariates, using greedy nearest neighbor matching with calipers of width equal to 0.1 SD of the logit of the propensity score. To assess covariate balance between the two study groups after matching, standardized mean differences (SMD) were calculated, with SMD<0.01 representing negligible covariate imbalance. The cumulative incidence of an outcome was calculated by dividing the number of patients in a group that had an occurrence of the outcome within the follow-up period by the number of patients in the group at the start of follow-up. Absolute risk differences (ARD) were calculated along with respective 95% confidence intervals (CI) as the difference between the cumulative incidence of an outcome in the tirzepatide group and the cumulative incidence of that outcome in the semaglutide group. Kaplan Meier curves were constructed with intergroup differences assessed using log-rank tests. Cox proportional hazards analysis was performed to calculate hazard ratios (HR) with 95% CIs.

Differences in treatment effects were estimated by performing subgroup analyses by age (65-years or older vs. Younger than 65 years), sex (male vs. Female), T2D, hypertension, chronic kidney disease (CKD), sleep apnea, ischemic heart disease (IHD), and atrial fibrillation. Additional propensity score matched cohorts were built for each subgroup analysis. The difference in treatment effects between patients who received tirzepatide and those who received semaglutide on the primary outcomes in each subgroup was analyzed, and the HR was calculated with the corresponding 95% CIs.

All analyses were conducted using the TriNetX platform (TriNetX LLC, Cambridge, MA, USA). A two-sided p value < 0.05 was considered statistically significant.

## RESULTS

### Study population

A total of 7,744,734 patients with obesity were identified in the TriNetX platform during the period Jun 4, 2021 to Aug 13, 2026. Of these, 1,049,511 patients met the exclusion and inclusion criteria and were initiated on tirzepatide or semaglutide therapy, with 431,938 using tirzepatide and 617,573 taking semaglutide. After propensity score matching, two equally-sized cohorts, with 424,269 patients in each, were formed (**Fig 1**).

**Fig 1.**
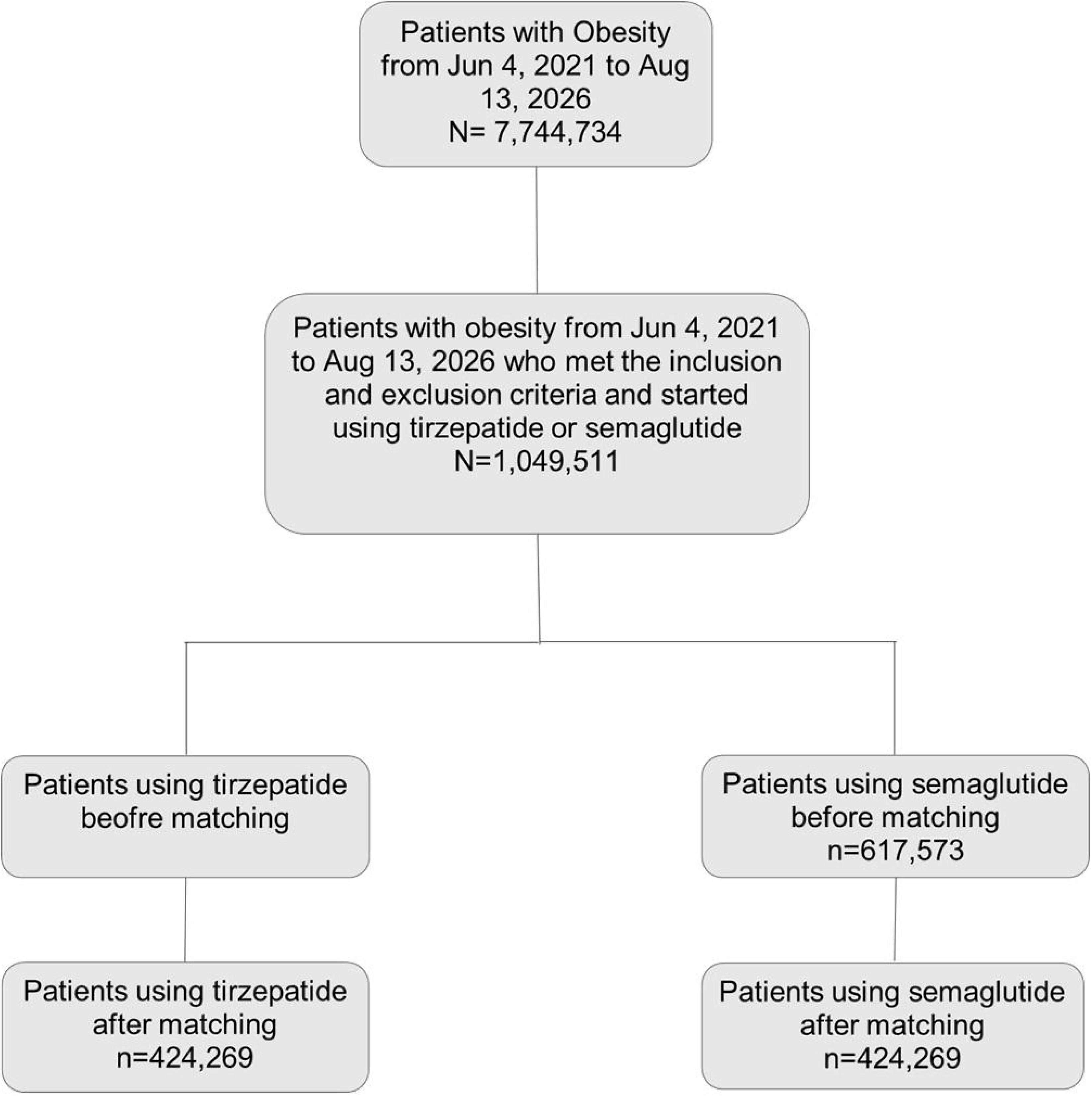
Study population flow diagram.

### Baseline characteristics

Baseline sociodemographic and clinical characteristics for the study groups before and after matching are reported in **Table 1**.

**Table 1.**
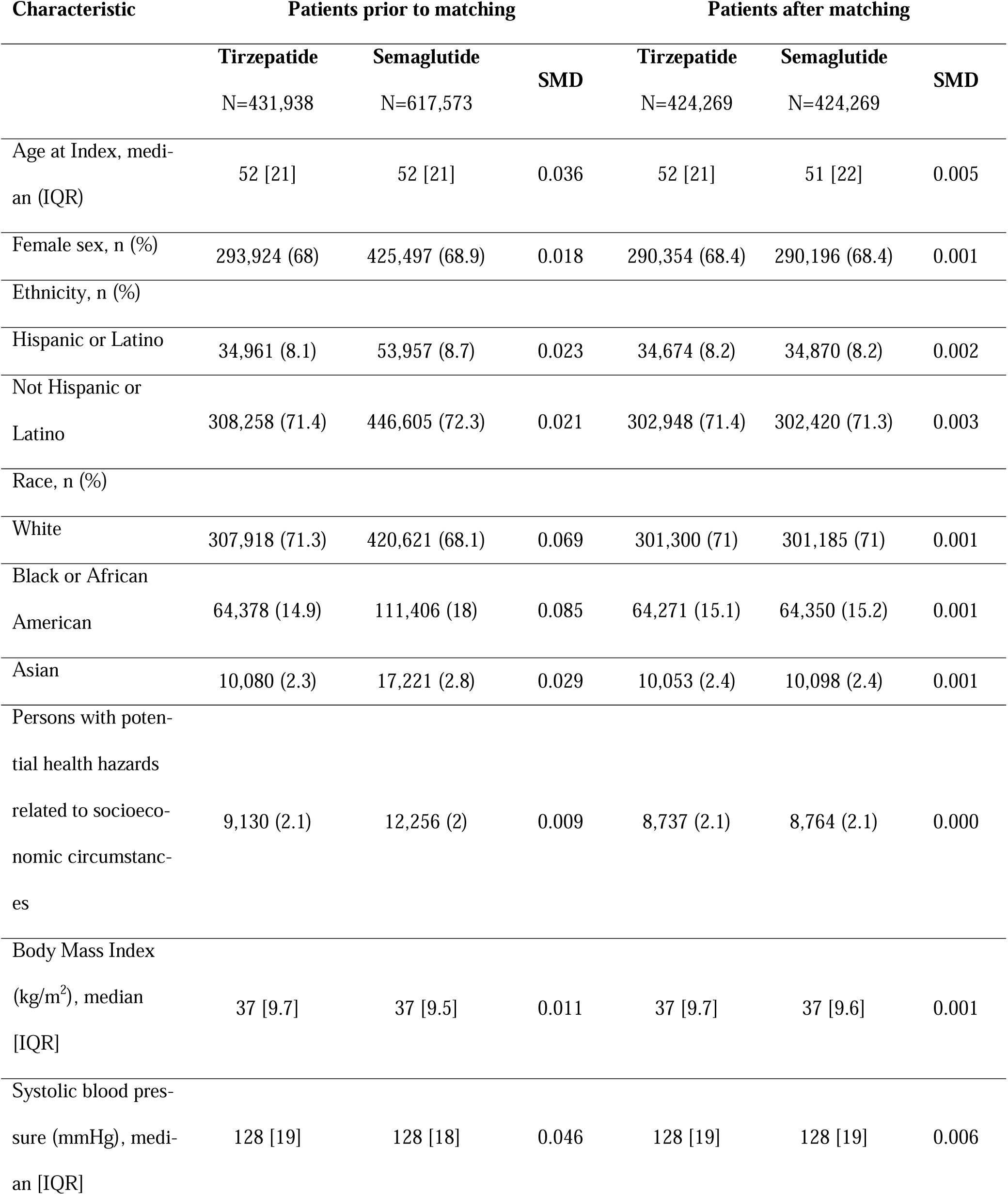
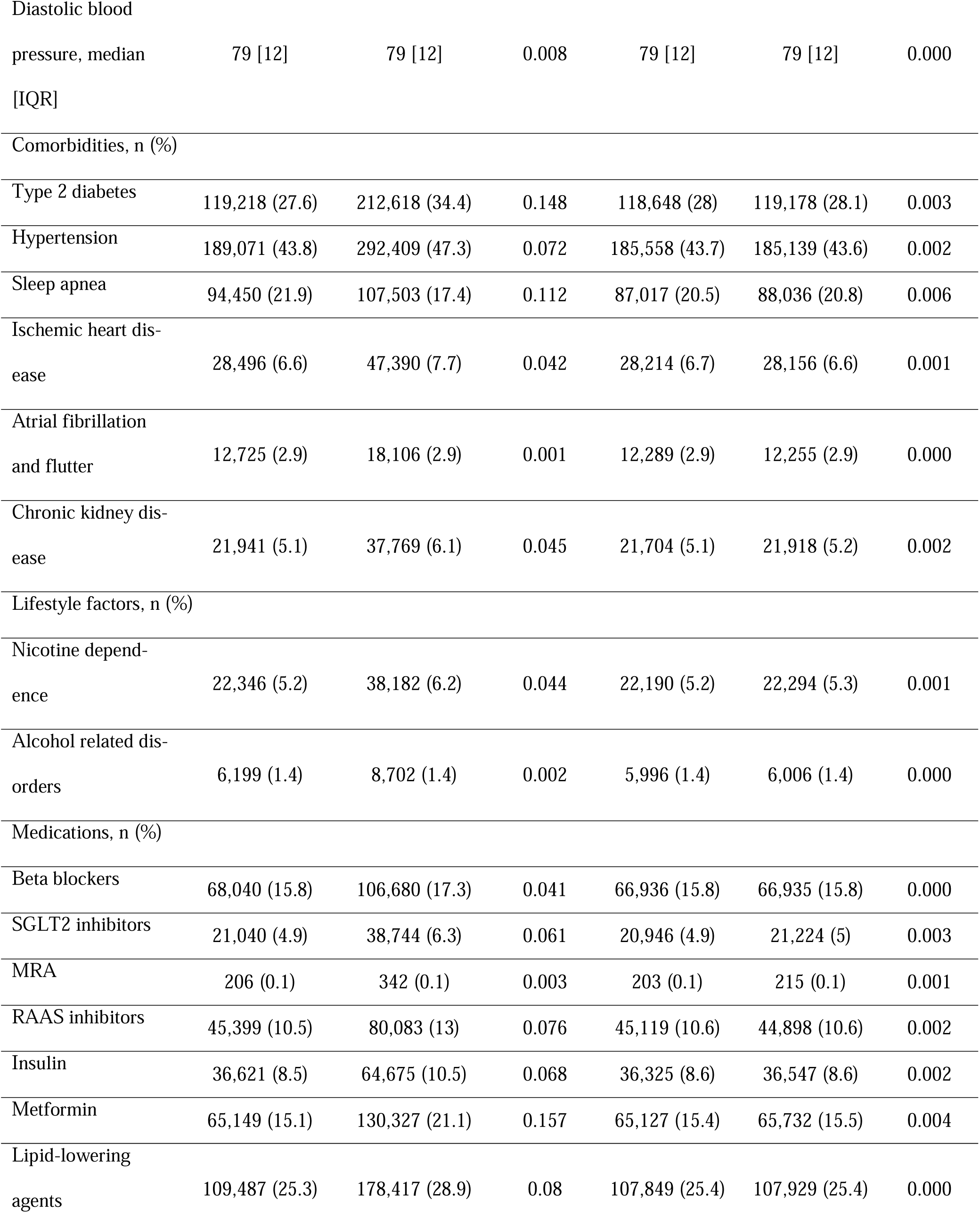
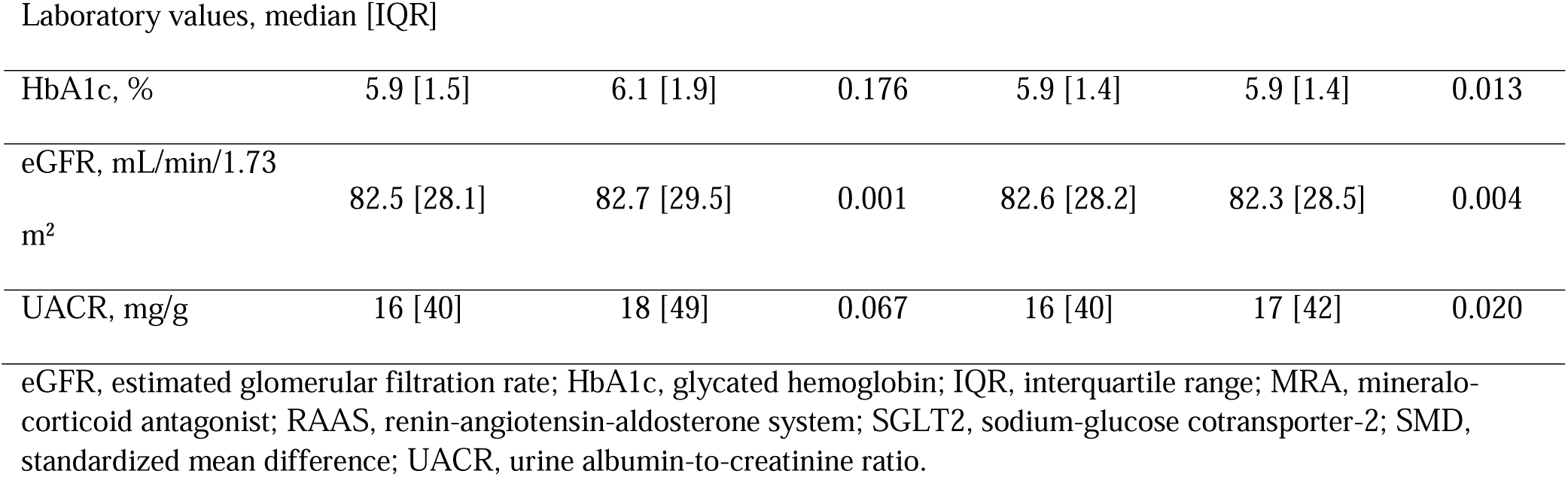
Baseline characteristics of patients with obesity who use tirzepatide or semaglutide.

In brief, before matching, both the tirzepatide and semaglutide groups had a median age of 52-years, and were predominantly female. The semaglutide group had more frequent diabetes compared to the tirzepatide group, whereas sleep apnea was more prevalent in the tirzepatide group. After propensity score matching, both groups had similar characteristics with SMD <0.1 for all covariates.

### Follow-up time

The median [interquartile range] follow-up time was 322 [238] days in the tirzepatide group and 365 [94] days in the semaglutide group. The majority of patients in both groups completed one year of follow-up (**Fig 2**).

**Fig 2.**
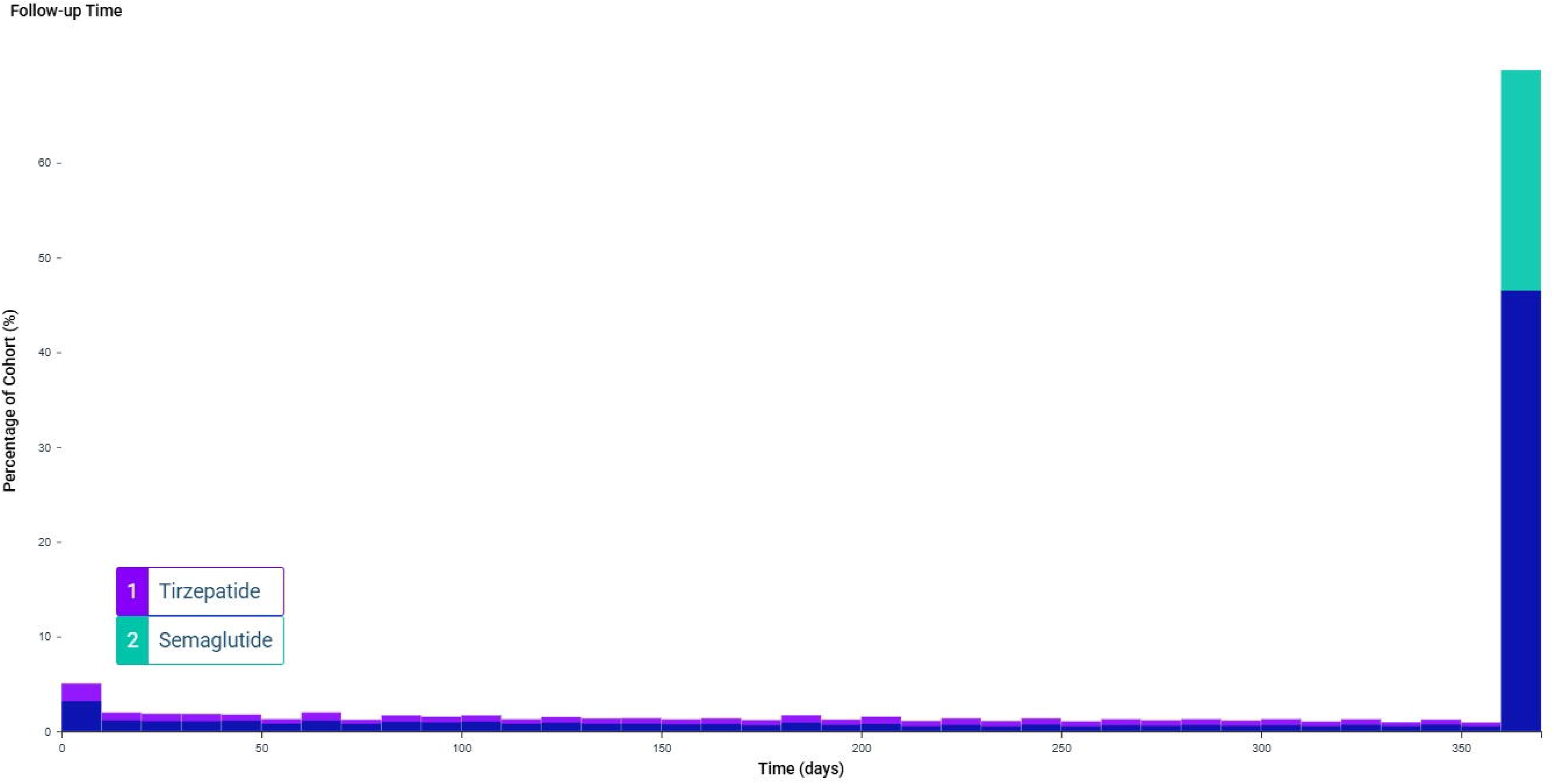
Follow-up time for patients with obesity using tirzepatide or semaglutide.

### Propensity score overlap

Propensity score overlap in the unmatched and matched cohorts are presented in **Fig 3**. The matched cohorts demonstrated good overlap.

**Fig 3.**
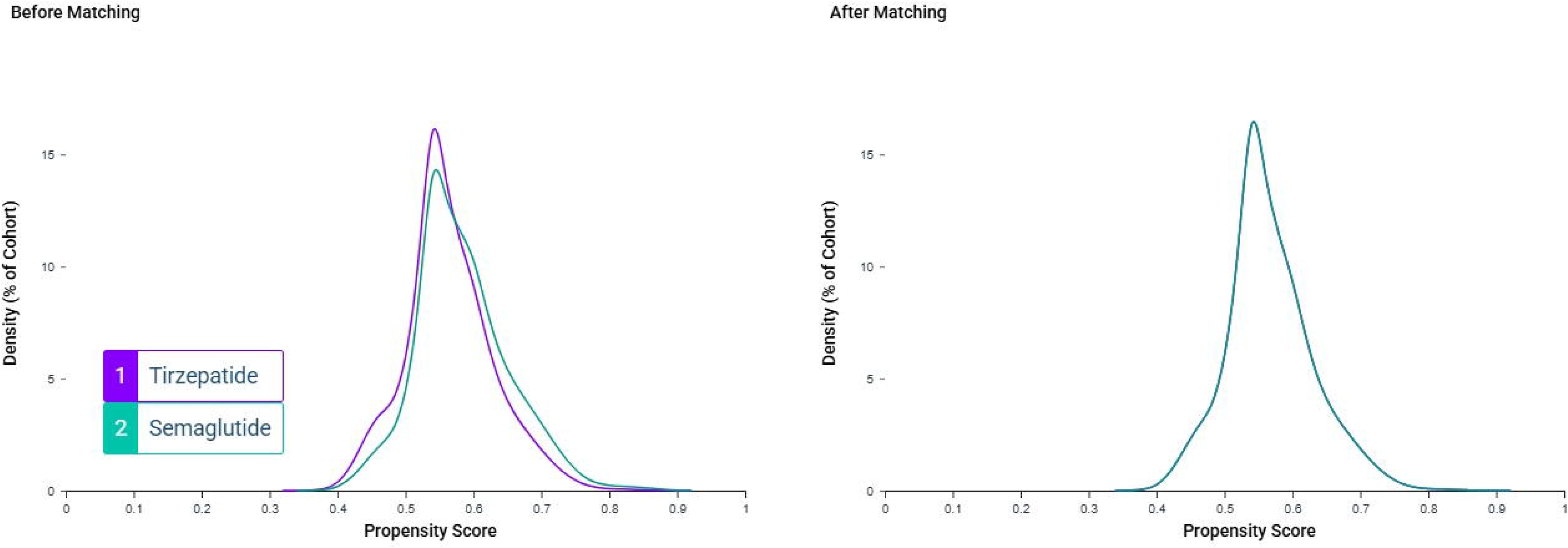
Propensity score density function of the unmatched and matched cohorts.

### Clinical outcomes

The ARDs and HRs, along with their respective 95% CIs, for the primary and secondary outcomes are reported in **Table 2**. Compared to the semaglutide group, the tirzepatide group had a lower incidence of NOHF, although the ARD was modest (−0.3%, 95% CI −0.35 to −0.26) (**Fig 4**). The tirzepatide group were also at lower risk of new-onset systolic HF and all-cause mortality compared to the semaglutide group. All other secondary outcomes occurred at similar rates in the two study groups.

**Fig 4.**
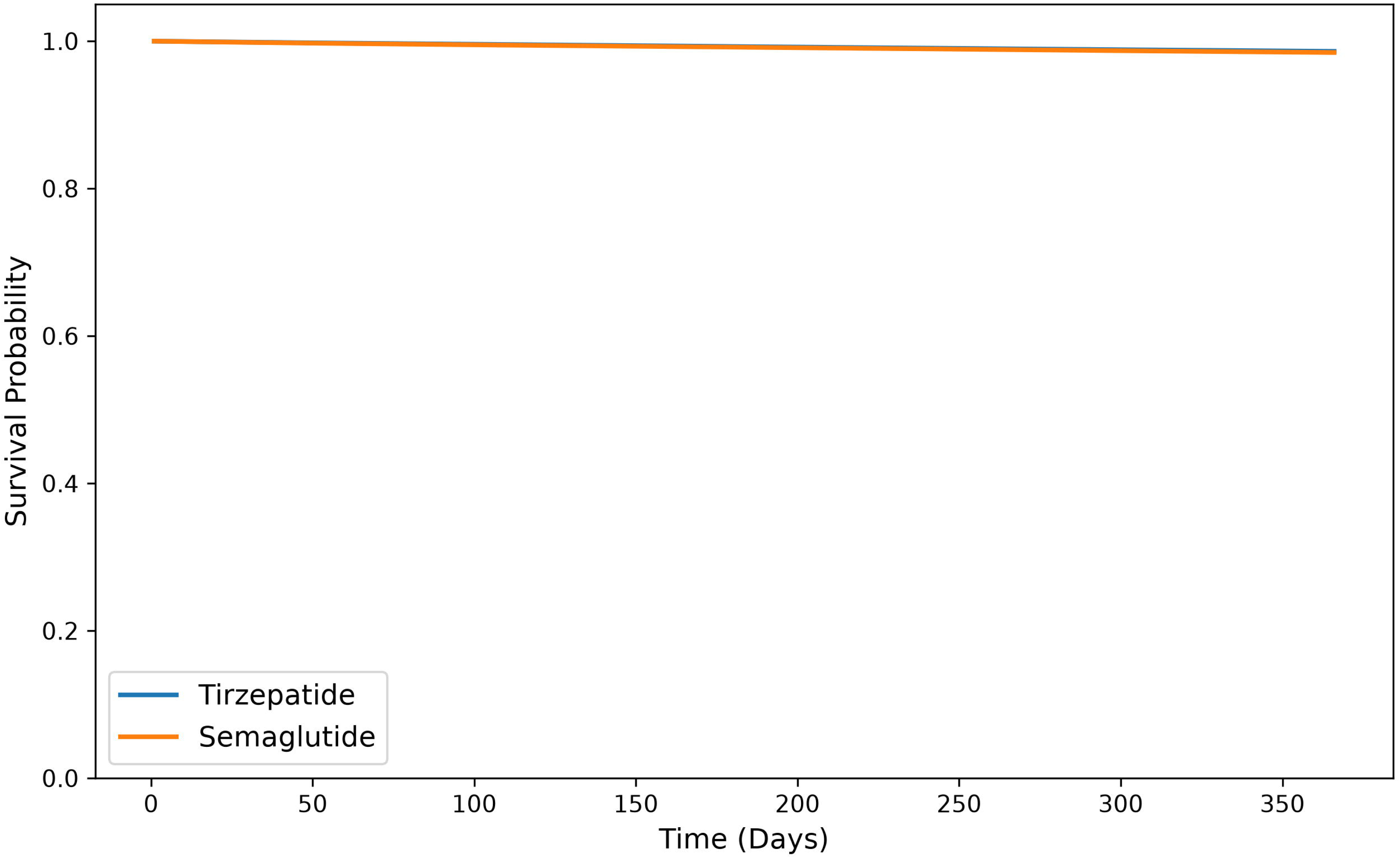
Kaplan-Meier curves comparing the incidence of new-onset heart failure between patients with obesity using tirzepatide or semaglutide. The online tool, TriNetX Publication Toolkit (URL: https://trinetxpublicationtoolkit.streamlit.app/) was accessed to generate Kaplan–Meier curves.

**Table 2.**
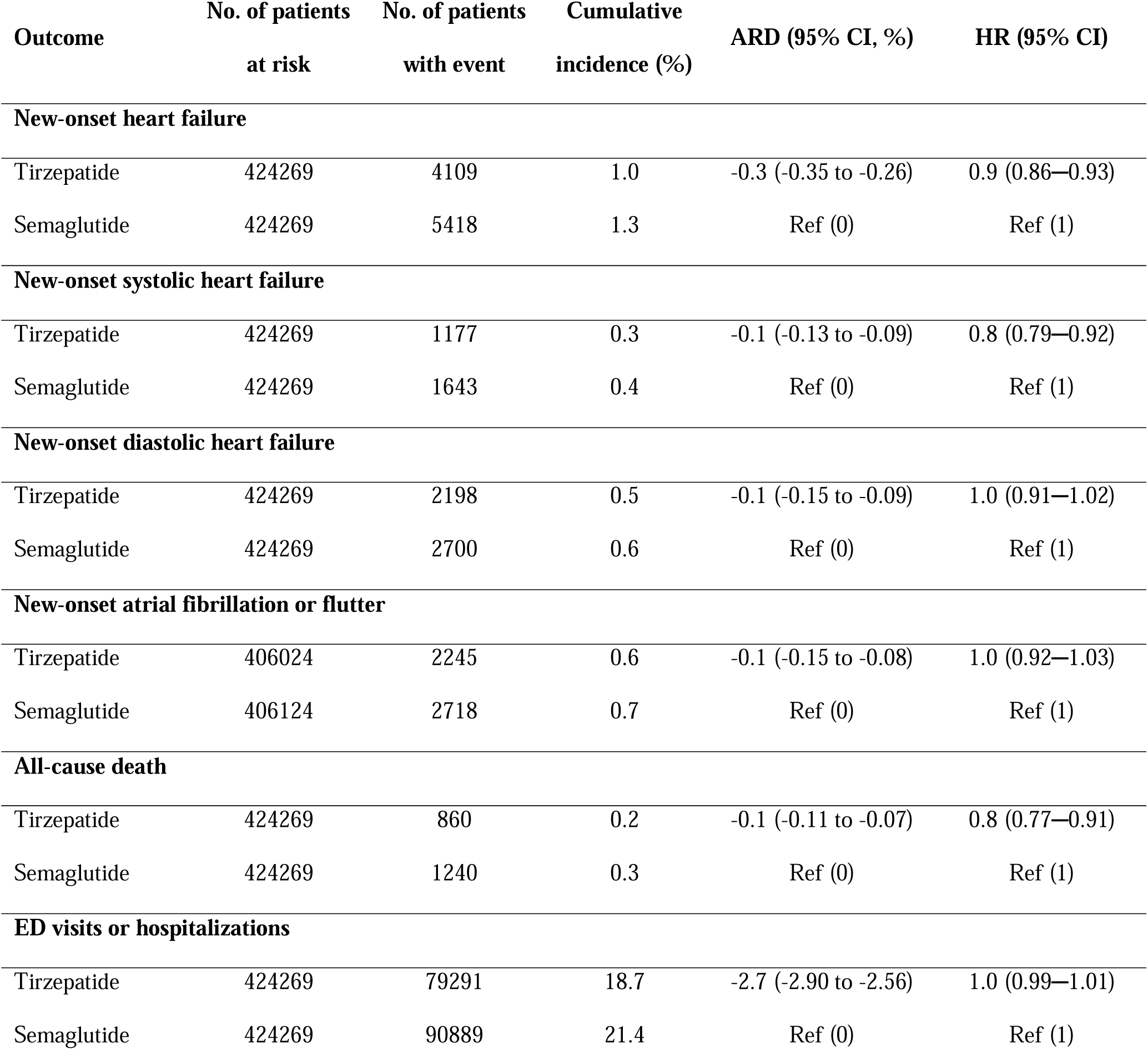
Incidence and hazard ratios of primary and secondary outcomes in patients with obesity treated with tirzepatide or semaglutide.

### Subgroup analysis

The incidence of NOHF was lower in the tirzepatide cohort compared to the semaglutide cohort of most analyzed subgroups of patients (**Fig 5**). Among patients with T2D, CKD, ischemic heart disease, atrial fibrillation, and patients without hypertension, there was no significant difference in the risk of NOHF between the respective tirzepatide and semaglutide cohorts.

**Fig 5.**
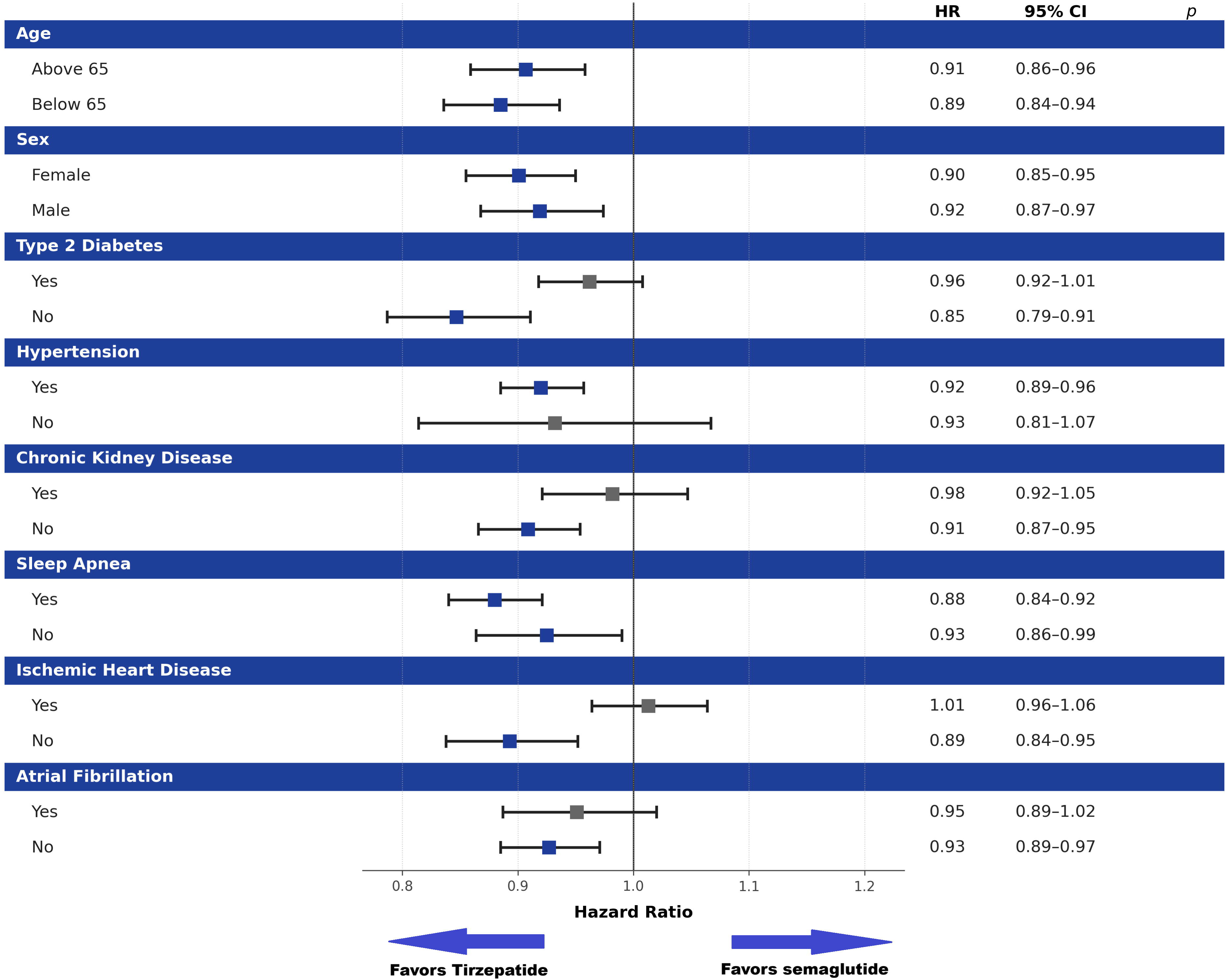
Forest plots presenting hazard ratios with 95% confidence intervals for new-onset heart failure in subgroups of patients with obesity using tirzepatide or semaglutide (reference). The online tool, TriNetX Publication Toolkit (URL: https://trinetxpublicationtoolkit.streamlit.app/) was accessed to generate forest plots.

## DISCUSSION

The current study employed a retrospective cohort design to explore and compare the incidence of NOHF among patients with obesity who were using tirzepatide or semaglutide. Patients who were prescribed tirzepatide during the study period had a lower incidence of NOHF compared to patients that used semaglutide although the ARD was modest at −0.3%. This trend was maintained in most analyzed subgroups of patients. Patients who were using tirzepatide were also at reduced risk of new-onset systolic HF and all-cause mortality compared to those who used semaglutide while other secondary outcomes occurred at similar rates in the two study groups.

Tirzepatide has been demonstrated to have greater efficacy in weight reduction and glycemic control compared to semaglutide in patients with obesity and T2D [2,10]. In the SUR-MOUNT-5 trial [2], patients with obesity who were randomized to the tirzepatide arm achieved greater weight reduction and control of glycated hemoglobin compared to the semaglutide arm. The tirzepatide group also reached greater control of other cardiometabolic risk factors including blood pressure, lipid profile and waist circumference. Similarly, in the SURPASS-2 trial [10], patients with T2D who took tirzepatide reached better control of body weight, HbA1c, blood pressure and lipid profile. Better control of cardiometabolic risk factors may play a central role in the reduced incidence of NOHF in patients treated with tirzepatide observed in the current study, especially in the case of HFpEF, where obesity is thought to be central to the etiopathogenesis of its common phenotype [5,26]. Several theories have been proposed to explain the increased benefit of dual agonism of GIP and GLP-1 receptors over GLP-1 agonism alone. Variable expression of GIP and GLP-1 receptors in the central nervous system may provide added neural targets for dual GIP/GLP1-RAs [1]. GIP signaling can also potentially modify adipose tissue function and promote healthy storage of dietary lipids by white adipose tissue as GIP receptors but not GLP-1 receptors are expressed in adipose tissue [31,33]. Adverse effects and drug discontinuation rates differed between tirzepatide and semaglutide in the SURMOUNT-5 and SURPASS-2 trials [2,10]. In the SURMOUNT-5 trial, the semaglutide arm experienced more side effects and had greater discontinuation rates compared to the tirzepatide arm [2]. Yet, in SURPASS-2, tirzepatide was associate with worse adverse event and discontinuation profiles compared to semaglutide although the absolute differences were modest in both trials [10]. In a recent systematic review and meta-analysis of randomized trials, both tirzepatide and semaglutide were associated with greater gastrointestinal side effects and drug discontinuation as compared with placebo [16]. However, there were no significant differences between the two drugs. In the SURPASS-CVOT trial [24], tirzepatide was non-inferior to dulaglutide in decreasing the risk of cardiovascular outcomes among patients with T2D. In a post-hoc analysis of SURPASS-CVOT [25], tirzepatide demonstrated greater reduction of cardiovascular and renal outcomes in patients with T2D. In a retrospective analysis of Medicare beneficiaries [21], tirzepatide and semagultide demonstrated similar efficacy in cardiovascular outcome risk reduction among patients with T2D. In a recent retrospective study comparing a broad range of cardiovascular outcomes between patients with T2D treated with tirzepatide or GLP1-RAs, therapy with tirzepatide was associated with lower risk of cardiovascular outcomes including HF exacerbations and new-onset systolic HF [8]. In the current study, tirzepatide was associated with a lower risk of NOHF compared to semaglutide in patients with obesity although the effect sizes were modest.

### Limitations

The current study adds to the existing literature on the differential effects of dual GIP/GLP1-RAs and GLP1-RAs on cardiovascular risk reduction, and provides evidence of lower risk of NOHF in patients with obesity who are treated with tirzepatide compared to those treated with semaglutide. The study is not without limitations however. The study used ICD-10 codes documented in the EHR of participating centers to define diagnoses and outcomes, which may have allowed for misclassification bias. Although the study performed propensity score matching along multiple covariates to limit the effect of confounding, other possible confounding factors may have not been accounted for due to the observational nature of the study. Other limitations inherent to observational study designs are also possible sources of bias.

## CONCLUSION

In a real-world analysis, patients with obesity who were treated with tirzepatide had lower risk of incident NOHF compared to matched patients treated with semaglutide although absolute risk differences were modest. Future randomized clinical trials and prospective studies are needed to validate and further explore these findings.

## Statements and Declarations

### Disclosure of potential conflicts of interest

The authors have no relevant financial or non-financial interests to disclose. Ethics approval This study was performed in line with the principles of the Declaration of Helsinki. All data used were collected from the TriNetX platform, which contains de-identified patient data only. Thus, an institutional review board (IRB) approval and patient consent were not required.

### Consent to participate declaration

The current study is exempt from informed consent. All data analyses are secondary analyses of existing data. The study does not involve intervention or interaction with human subjects. All included data are de-identified per the de-identification standard defined in Section §164.514(a) of the HIPAA Privacy Rule.

### Funding declaration

This research did not receive any specific grant from funding agencies in the public, commercial, or not-for-profit sectors.

### Declaration of generative AI and AI-assisted technologies in the manuscript preparation process

During the preparation of this work the authors used the online tool, TriNetX Publication Toolkit (URL: https://trinetxpublicationtoolkit.streamlit.app/) in order to generate Kaplan–Meier curves and forest plots. After using this tool/service, the authors reviewed and edited the content as needed and takes full responsibility for the content of the published article.

### Author Contribution Statement

Conceptualization, M.A.; Investigation, M.A., T.A.H., S.K. and J.A.; Formal analysis, M.A.; Methodology, M.A., T.A.H., S.K. and J.A.; Project administration, M.A., S.K. and J.A.; Supervision, M.A. and J.A.; Validation, M.A. and J.A.; Visualization, M.A., S.K. and J.A.; Writing - original draft, M.A. and T.A.H.; Writing - review & editing, M.A., S.K. and J.A.

## Data Availability

All data produced are available online at (https://live.trinetx.com/).

## Notes

### Competing Interest Statement

The authors have declared no competing interest.

### Author Declarations

This retrospective study is exempt from Institutional Review Board (IRB) approval. The data reviewed is a secondary analysis of existing data that can be accessed online at the following link (https://live.trinetx.com/). The study does not involve intervention or interaction with human subjects, and is de-identified per the de-identification standard defined in Section 164.514(a) of the HIPAA Privacy Rule.

