## Supplementary Materials for "New-Onset Heart Failure in Patients with Obesity using Tirzepatide or Semaglutide"

**Table S1**. Covariate definitions

| **Covariate** | **Definition (ICD-10 Code)** |
| --- | --- |
| Socioeconomic status | Z55-Z65 |
| hypertension | I10 |
| dyslipidemia | E78 |
| ischemic heart disease | I20-I25 |
| atrial fibrillation or flutter | I48 |
| valvular heart disease | I05-I08; I34-I37 |
| cerebrovascular disease | I60-I69 |
| peripheral arterial disease | I70-I79 |
| sleep apnea | G47.3 |
| Chronic kidney disease | N18 |
| chronic obstructive pulmonary disease | J44 |
| liver disease | K70-K77 |
| tobacco smoking | F17 |
| alcohol use disorder | F10 |
| Insulin | ATC- A10A |
| Metformin | RXCUI- 6809 |
| Sodium-glucose cotransporter 2 inhibitors | ATC- A10BK |
| Lipid-lowering medications | ATC- C10 |
| beta-blockers | ATC- C07 |
| renin-angiotensin-aldosterone system inhibitors | ATC- C09 |
| Mineralocorticoid receptor antagonists | ATC- C03DA |
| Anti-hypertensive | ATC- C02 |
